# Circulating insulin-like growth factor-I, total and free testosterone concentrations and prostate cancer risk in 200,000 men in UK Biobank

**DOI:** 10.1101/2020.03.27.20044941

**Authors:** Eleanor L. Watts, Georgina K. Fensom, Karl Smith Byrne, Aurora Perez-Cornago, Naomi E. Allen, Anika Knuppel, Marc J. Gunter, Michael V. Holmes, Richard M. Martin, Neil Murphy, Konstantinos K. Tsilidis, Bu B. Yeap, Timothy J. Key, Ruth C. Travis

## Abstract

**Background:** Insulin-like growth factor-I (IGF-I) and testosterone have been implicated in prostate cancer aetiology. Using newly available data from a large prospective full-cohort with standardised assays and repeat blood measurements, and genetic data from an international consortium, we aimed to investigate the associations of circulating concentrations of IGF-I, sex hormone-binding globulin (SHBG), total and calculated free testosterone with prostate cancer risk.

**Patients and methods:** For prospective analyses of prostate cancer incidence and mortality, we studied 199,698 male UK Biobank participants using Cox proportional hazards models. Multivariable-adjusted hazard ratios (HRs) were corrected for regression dilution bias using repeat hormone measurements from a subsample of up to 7,776 men. A 2-sample Mendelian randomization (MR) analysis of IGF-I and risk used genetic instruments identified from UK Biobank men and genetic outcome data from 79,148 cases and 61,106 controls from the PRACTICAL consortium. We used *cis*- and all (*cis* and *trans*) SNP MR approaches.

**Results:** After a mean follow-up of 6.9 years, 5,402 men were diagnosed with and 295 died from prostate cancer. Higher circulating IGF-I was associated with an elevated risk (HR per 5 nmol/L increment=1.09, 95% CI 1.05-1.12) and prostate cancer mortality (HR per 5 nmol/L increment=1.15,1.02-1.29) in observational analyses. *Cis*- and all SNPs MR analyses also supported the role of IGF-I in prostate cancer diagnosis (*cis*-MR odds ratio per 5 nmol/L increment=1.34,1.07-1.68). In observational analyses, higher free testosterone was associated with a higher risk of prostate cancer (HR per 50 pmol/L increment=1.10,1.05-1.15), and higher SHBG was associated with a lower risk of prostate cancer (HR per 10 nmol/L increment=0.95,0.94-0.97), but neither was associated with prostate cancer mortality. Total testosterone was not associated with prostate cancer.

**Conclusion(s):** These findings implicate IGF-I and free testosterone in prostate cancer development and/or progression.

**Key message:** Prostate cancer is the second most common cancer in men, but modifiable risk factors have not been identified. Using large-scale prospective observational and genetic data we investigated the associations of circulating IGF-I, SHBG and testosterone with prostate cancer diagnosis and mortality. Our results implicate IGF-I and the sex hormones in prostate cancer development.

**Highlights:**

- Prostate cancer is the second most common cancer in men, but no modifiable risk factors are established.
- We examined the associations of circulating IGF-I, SHBG, total and free testosterone with prostate cancer risk in UK Biobank.
- Men with higher IGF-I had a higher risk of diagnosis and mortality. Mendelian randomization also supported a causal role.
- Men with higher free testosterone had a higher prostate cancer risk, and men with higher SHBG had a lower risk.
- The results implicate IGF-I and the sex hormones in prostate cancer development.

## Introduction

Prostate cancer is the second most common cancer in men worldwide, and a leading cause of cancer death^1^. Few potential modifiable risk factors have been identified, but circulating hormone concentrations are thought to play a role in prostate cancer aetiology^2 3^.

Insulin-like growth factor-I (IGF-I) is involved in cell proliferation, differentiation and apoptosis, and prospective studies have shown a positive association of circulating IGF-I concentration with prostate cancer risk^4^. Less is known about its potential role in prostate cancer progression or mortality^5^.

Androgens are integral to the maintenance of normal prostate function^6^. In the circulation, testosterone is bound to sex hormone-binding globulin (SHBG) and albumin. Approximately 2% of total testosterone circulates unbound or “free” and is postulated to be biologically active^7^. Observational pooled analysis of individual participant data from prospective studies indicated that men with very low free testosterone may have a lower risk of prostate cancer^8^, and a recent Mendelian randomization (MR) study supports a positive association between free testosterone concentration and prostate cancer diagnosis^9^. However, it is unclear whether circulating free testosterone concentration is associated with prostate cancer mortality^8 10^. Epidemiological studies have also reported an inverse association between prostate cancer risk and circulating SHBG^8^, although results from MR analyses are inconclusive^9^.

Previous risk estimates for prostate cancer in relation to hormone concentration have generally been based on data from nested case-control studies with a single blood draw at baseline. The UK Biobank study has standardised measurements of hormones from baseline blood samples collected in the whole cohort (500,000 participants) as well as repeat measurements of the hormones in a subset (20,000).

In this paper, we aimed to examine the associations of serum concentrations of IGF-I, SHBG, total and free testosterone with prostate cancer incidence and mortality, using observational data from UK Biobank. For IGF-I, we investigated potential causal associations of IGF-I with prostate cancer using MR analyses, with genetic data from UK Biobank and the PRACTICAL consortium (based on 79,000 prostate cancer cases and 61,000 controls). MR analyses of SHBG, total and free testosterone and prostate cancer risk have recently been published^9^. MR uses germline genetic variants as proxies of putative risk factors and estimates their associations with disease. As germline genetic variants are fixed and randomly allocated at conception, this technique minimizes the chance of confounding and reverse causality, and is therefore considered a useful approach towards causal inference^11^. By using these two complementary approaches we were able to robustly investigate associations and assess causation.

## Patients and methods

### UK Biobank-observational analysis

#### Study design

UK Biobank is a prospective cohort with open-access for public health research. Details of the study protocol and data collection are available online (http://www.ukbiobank.ac.uk/wp-content/uploads/2011/11/UK-Biobank-Protocol.pdf) and elsewhere^12 13^.

In brief, all participants were registered with the UK National Health Service (NHS) and lived within 40 km of one of the UK Biobank assessment centres. Approximately 9.2 million people were initially invited to participate. Overall, 503,317 men and women aged 40-69 years consented to join the cohort and attended one of 22 assessment centres throughout England, Wales and Scotland between 2006-2010, a participation rate of 5.5%^13^.

The UK Biobank study was approved by the North West Multi-Centre Research Ethics Committee (reference number 06/MRE08/65), and at recruitment all participants gave informed consent to participate and for their health to be followed-up through linkage to electronic medical records.

#### Baseline assessment

At the baseline assessment visit, participants provided information on a range of sociodemographic, physical, lifestyle, and health-related factors via a self-completed touch-screen questionnaire and a computer assisted personal interview^13^. Weight and height were measured at the assessment centre^13^.

#### Blood sampling and biomarker assays

At recruitment, blood sampling was successfully performed in 99.7% of the cohort. Blood was collected in a serum separator tube and shipped to the central processing laboratory in temperature-controlled boxes at 4°C^14^, then aliquoted and stored in a central working archive at −80°C^15^. Serum concentrations of circulating IGF-I, SHBG, testosterone and albumin were measured in all participants. IGF-I (DiaSorin Liaison XL), SHBG and testosterone (Beckman Coulter AU5800) were determined by chemiluminescent immunoassays. Albumin was measured by a colourimetric assay (Beckman Coulter AU5800). Average within-laboratory (total) coefficients of variation for low, medium, and high internal quality control level samples for each biomarker ranged from 2.1-8.3%. Full details of the assay methods and quality assurance protocols are available online (https://biobank.ndph.ox.ac.uk/showcase/docs/serum_biochemistry.pdf).

#### Free testosterone estimation

Free testosterone concentrations were estimated using a formula based on the law of mass action from measured total testosterone, SHBG and albumin concentrations^8 16^.

#### Repeat assessment

Participants who lived within a 35 km radius were invited to attend a repeat assessment clinic at the UK Biobank Co-ordinating Centre in Stockport between August 2012 and June 2013. Repeat assessments were completed in 20,000 participants (9,000 men) with a response rate of 21%^17^.

#### Participant follow-up

Cancer registration data were provided via record linkage to the NHS Central Register and obtained via NHS Digital, until the censoring date (31^st^ March 2016 in England and Wales and 31^st^ October 2015 in Scotland). Death data for England and Wales were provided by NHS Digital and for Scotland by the Information and Statistics Division (censoring dates 31^st^ January 2018 in England and Wales, and 30^th^ November 2016 in Scotland). In the analysis of incident prostate cancer, the endpoint was defined as the first diagnosis of prostate cancer, or prostate cancer mortality (primary or otherwise) (International Classification of Diseases Tenth revision code [ICD-10] C61^18^), whichever was recorded first. In the analysis of prostate cancer mortality, the endpoint was prostate cancer as the primary cause of death. Person-years were calculated from the date of recruitment to the date of the first cancer registration (excluding non-melanoma skin cancer [ICD-10 C44]), death, or censoring date, whichever occurred first.

#### Exclusion criteria

Our analytical dataset included 199,698 men; we excluded 9,871 men with prevalent cancer (except C44: non-melanoma skin cancer), 13,509 men who did not have blood data available or who had biomarker measurements that did not pass quality control procedures^19^, 1,685 participants for whom it was not possible to determine genetic sex or who were identified as being genetically female, 2,326 men who reported taking hormone medication at baseline, and 758 men who had no body mass index (BMI) data.

#### Statistical analysis

Hazard ratios (HRs) and 95% confidence intervals (CIs) of prostate cancer diagnosis and mortality were estimated using Cox proportional hazards models, with age as the underlying time variable. Analyses were stratified by geographic area (10 UK regions) and age at recruitment (<45, 45-49, 50-54, 55-59, 60-64, ≥65 years), and adjusted for Townsend deprivation score (fifths, unknown (0.1%)), racial/ethnic group (white, mixed background, Asian, black, other, and unknown (0.5%)), height (<170, ≥170–<175, ≥175–<180, ≥180 cm, and unknown (0.1%)), lives with a wife or partner (no, yes), body mass index (BMI) (<25, ≥25–<30, ≥30–<35, ≥35 kg/m^2^), cigarette smoking (never, former, current light smoker (1-<15 cigarettes per day), current heavy smoker (≥15 cigarettes per day), current (number of cigarettes per day unknown), and smoking status unknown (0.6%)), alcohol consumption (non-drinkers, <1-<10, ≥10-<20, ≥20 g ethanol/day, unknown (0.5%)), and self-reported diabetes (no, yes, and unknown (0.5%)). Adjustment covariates were defined *a priori* based on previous analyses of UK Biobank data^20^.

Blood biomarker measurements were also available for up to 7,776 men who attended a repeat assessment clinic a median of 4.4 years after first blood collection^17^. Measurement error and within person variability using single measures at baseline leads to under-estimation of risk (i.e. regression dilution bias)^21^; to provide more precise and generalizable risk estimates, HRs for trend were estimated per absolute increase in usual hormone concentrations, with correction for regression dilution bias using the McMahon-Peto method^21 22^.

In the categorical analyses, biomarker measurements were categorised into fifths based on the distribution in the whole cohort and HRs were calculated relative to the lowest fifth of each blood parameter. The variance of the log risk in each group was calculated (from the variances and covariances of the log risk) and used to obtain group-specific 95% CIs, which enable comparisons across different exposure categories^23^.

The proportional hazards assumption was examined using time-varying covariates and Schoenfeld residuals, and revealed no evidence of deviation.

#### Subgroup analyses

Subgroup analyses for incident prostate cancer were examined by the following categories: age at diagnosis (≤65, >65 years), time from blood collection to diagnosis (≤4, >4 years), age at blood collection (<60, ≥60 years), BMI (<30, ≥30 kg/m^2^), smoking status (never or former, current), alcohol consumption (<10, ≥10 g ethanol/day), education status (no university degree, university degree), currently married/cohabiting (no, yes), Townsend index (<median, ≥median), ethnicity (white, non-white), height (≤175, >175 cm), diabetes (no, yes), family history of prostate cancer (no, yes), poor self-rated health (no, yes) and median observed hormone concentrations (<median, ≥median). Subgroup categories were chosen *a priori* on the basis of data distributions and previous analyses by this research group^4 8^. Heterogeneity in the associations for case-specific variables (i.e. age at diagnosis and time from blood collection to diagnosis) was examined using stratified Cox models based on competing risks and comparing the risk coefficients and standard errors in the two subgroups, and testing with a χ^2^ for heterogeneity. For non-case specific factors, heterogeneity was assessed using a χ^2^ interaction term. Heterogeneity in the associations with prostate cancer mortality was not tested due to the limited statistical power.

#### Further analyses

We examined the possible association of IGF-I with incident prostate cancer after additional adjustment for concentrations of free testosterone and SHBG. We also investigated the associations of total and free testosterone and SHBG after further adjustment for IGF-I (with adjustment for biomarkers categorised into fifths and unknown). As a further examination of trend, the categorical variable representing the fifths of the hormones was replaced with a continuous variable that was scored as 0, 0.25, 0.5, 0.75, and 1, such that a unit increase in this variable can be taken to represent an 80 percentile increase in concentrations to enable comparison across hormones and with previous pooled analyses^4 8^. Analyses with prostate cancer diagnosis were repeated after hormone concentrations were divided into tenths.

All analyses were performed using Stata version 14.1 (Stata Corporation, College Station, TX, USA), and figures were plotted in R version 3.2.3. All tests of significance were two-sided, and P-values <0.05 were considered statistically significant.

### Mendelian randomization analyses

#### Genetic associations with circulating IGF-I

Single nucleotide polymorphisms (SNPs) associated with circulating IGF-I (*P*<5 × 10^−8^ significance threshold) were identified from a publicly available GWAS from 167,174 male UK Biobank participants of European ancestry^24^. SNPs were pruned by a linkage disequilibrium (LD) threshold of r^2^ < 0.01. UK Biobank genotyping details are reported elsewhere^25^.

GWAS results were partitioned into one primary *cis-*SNP instrument within the IGF-I gene region on chromosome 12 (rs5742653), and 216 additional *trans-*SNPs (SNPs associated with circulating IGF-I concentrations that are not located in this gene region). These *cis* and *trans-* SNPs together explained 9.3% of the variance in circulating concentrations of IGF-I. SNP rs numbers, nearest gene, and effect estimates are displayed in Supplementary Table S1.

#### Genetic instruments for prostate cancer

We used summary statistics for SNP associations with prostate cancer risk that were generated from 79,148 prostate cancer cases and 61,106 controls of European ancestry from the PRACTICAL, CRUK, CAPS, BP3 and PEGASUS consortia^26 27^. In brief, 44,825 prostate cancer cases and 27,904 controls were genotyped using OncoArray (http://epi.grants.cancer.gov/oncoarray/), and data were also available from several previous prostate cancer GWAS: UK stage 1 and stage 2; CaPS 1 and CaPS 2; BPC3; NCI PEGASUS; and iCOGS. Genotype information was imputed for all samples using the October 2014 release of the 1000 Genomes Project data as the reference panel. Odds ratios (ORs) and standard errors were estimated using logistic regression and then meta-analysed using an inverse variance fixed effect approach.

Where valid SNPs identified in UK Biobank were not present in PRACTICAL, we used HaploReg^28^ to identify SNPs in linkage disequilibrium (r^2^□>□0.8) to use as proxies.

#### Statistical analysis

We used a 2-sample MR approach to estimate IGF-I associations with overall prostate cancer risk, using UK Biobank as our genetic instruments for IGF-I and PRACTICAL for genetic outcome analyses.

The MR estimation for IGF-I and prostate cancer was conducted by the Wald ratio using the *cis*-SNP (rs5742653). We also conducted analyses incorporating all 217 IGF-I associated SNPs using the inverse-variance weighted method, as well as weighted median and mode-based methods to reduce the influence of pleiotropy^31^. To further assess the potential presence of horizontal pleiotropy we used Cochran’s Q for heterogeneity^31^ and the intercept from the MR-Egger method. Additionally, we used leave-one-out analyses to test the sensitivity of our results to single SNP effects. PhenoScanner was used to assess pleiotropy of the genetic instruments^32 33^.

Statistical analyses were performed using the *TwoSampleMR* R package^34^.

## Results

### UK Biobank observational analyses

After a mean follow-up of 6.9 years (standard deviation [SD]=1.3 years), 5,402 (2.7%) men were diagnosed with prostate cancer and 297 died from the disease. Table 1 summarises the baseline characteristics of study participants. Mean age at recruitment was 56.5 years (SD=8.2), and mean BMI was 27.8 kg/m^2^. 29% reported having had a PSA test prior to baseline and 14% had a family history of prostate cancer. Means and SDs for baseline biomarker measurements are displayed in Table 1. Regression dilution ratios ranged between 0.57 (free testosterone) and 0.80 (IGF-I) (Supplementary Table S2).

**Table 1:** Baseline characteristics and blood data for all men and for men who developed prostate cancer in UK Biobank.

|  | All men (*N*=199,698) | Men who developed prostate cancer (*N*=5,402) |
| --- | --- | --- |
| **Sociodemographic** |  |  |
| Age at recruitment (years), mean (SD) | 56.5 (8.19) | 62.1 (5.26) |
| Most deprived quintile, % (*N*) | 19.7 (39280) | 15.8 (854) |
| Black ethnicity, % (*N*) | 1.45 (2882) | 2.18 (117) |
| Not in paid/self-employment, % (*N*) | 38.3 (76448) | 56.7 (3062) |
| Living with partner, % (*N*) | 92.9 (152616) | 95.5 (4281) |
| **Anthropometric, mean (SD)** |  |  |
| Height (cm) | 175.7 (6.84) | 175.1 (6.68) |
| BMI (kg/m<sup>2</sup>) | 27.8 (4.23) | 27.5 (3.79) |
| **Lifestyle, % (*N*)** |  |  |
| Current cigarette smokers | 12.5 (24855) | 9.35 (502) |
| Drinking alcohol ≥20 g per day | 43.7 (86905) | 42.7 (2298) |
| Low physical activity (0-10 METs per week) | 28.1 (54408) | 26.6 (1393) |
| **Health history, % (*N*)** |  |  |
| Hypertension | 52.2 (104099) | 58.6 (3163) |
| Diabetes | 6.82 (13545) | 5.91 (318) |
| Poor self-rated health | 4.81 (9547) | 3.14 (169) |
| **Prostate specific factors, % (*N*)** |  |  |
| Ever had a PSA test | 28.7 (54139) | 46.5 (2396) |
| Family history of prostate cancer | 13.6 (14925) | 25.0 (703) |
| **Baseline blood measures, mean (SD)** |  |  |
| IGF-I (nmol/L) | 21.9 (5.52) | 21.6 (5.23) |
| SHBG (nmol/L) | 39.5 (16.6) | 41.9 (16.0) |
| Total testosterone (nmol/L) | 12.0 (3.65) | 12.0 (3.53) |
| Free testosterone (pmol/L) | 209 (59.5) | 200 (54.5) |
| Albumin (g/L) | 45.6 (2.61) | 45.2 (2.53) |

Abbreviations: BMI=body mass index; IGF-I=insulin-like growth factor-I; METs=metabolic equivalent of tasks; PSA=prostate-specific antigen; SD=standard deviation; SHBG=sex hormone binding globulin.

#### Associations between hormone concentrations and prostate cancer risk

Serum IGF-I concentration was positively associated with prostate cancer incidence (HR per 5 nmol/L increment=1.09, 95% CI 1.05-1.12; *P*_*trend*_<0.0001, Figure 1) and prostate cancer mortality (HR per 5 nmol/L increment =1.15, 95% CI 1.02-1.29; *P*_*trend*_=0.03, Figure 2).

**Figure 1:**
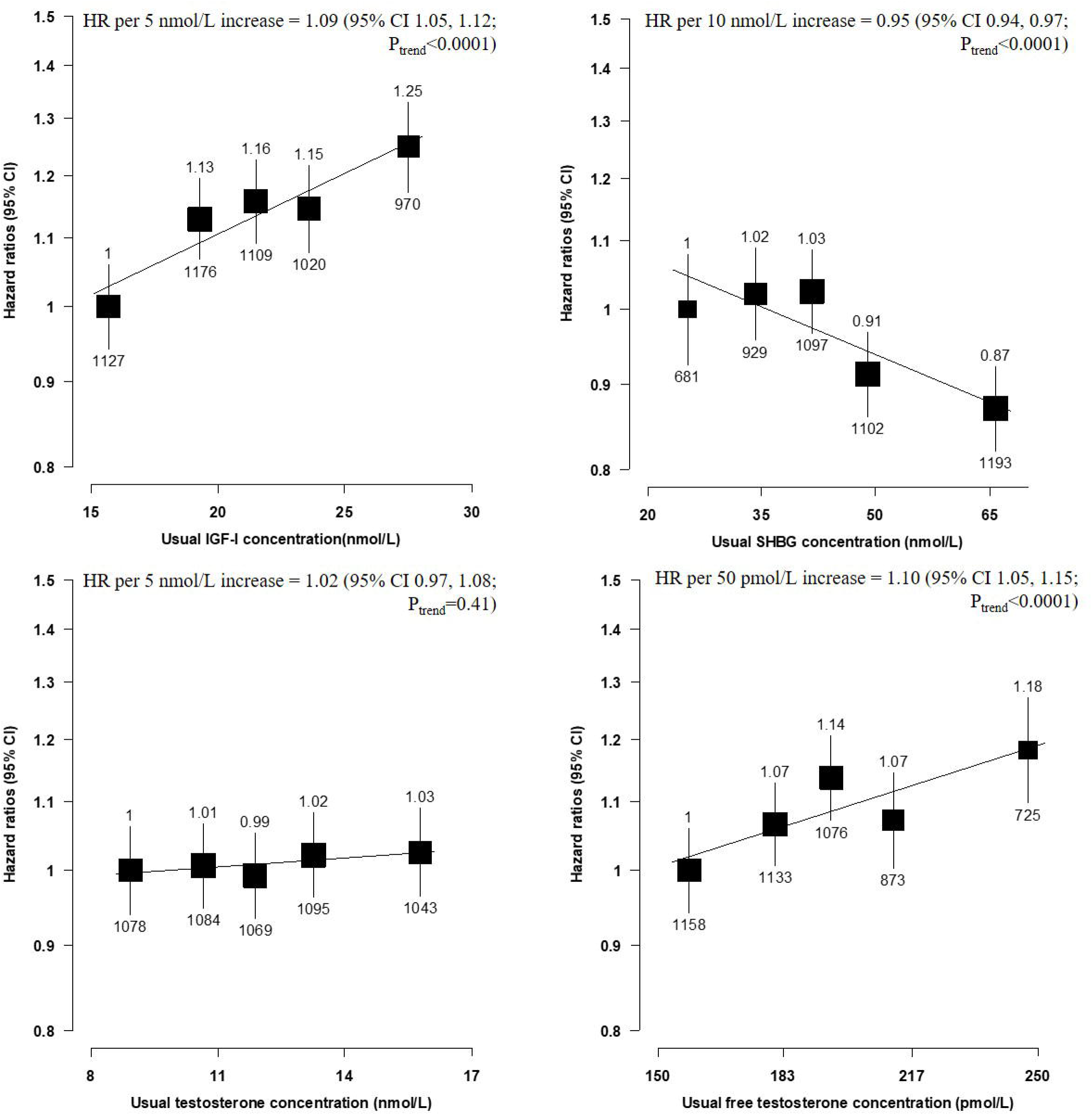
Hazard ratios of incident prostate cancer by fifths of usual serum hormone concentrations in UK Biobank. HRs are stratified by region (10 UK cancer registry regions) and age at recruitment (<45, 45–49, 50–54, 55–59, 60–64, and ≥65 years) and adjusted for age (underlying time variable), Townsend deprivation score (fifths, unknown), racial/ethnic group (white, mixed background, Asian, black, other, unknown), height (<170, ≥170– <175, ≥175–<180, ≥180 cm, unknown), lives with a wife or partner (no, yes), BMI (<25, ≥25–<30, ≥30–<35, ≥35 kg/m^2^), cigarette smoking (never, former, light smoker, heavy smoker, current unknown, and smoking status unknown), alcohol consumption (non-drinkers, <1-<10, ≥10-<20, ≥20 g ethanol/day, unknown), and diabetes (no, yes, and unknown). HRs for trend are adjusted for regression dilution bias. The boxes represent the HRs; the vertical lines represent the 95% CIs, with the size inversely proportional to the variance of the logarithm of the HR. The numbers above the vertical lines are point estimates for HRs, and the numbers below are the number of prostate cancer diagnoses. Abbreviations: BMI= body mass index; CI= confidence intervals; HR= hazard ratio; IGF-I=insulin-like growth factor-I; SHBG=sex hormone binding globulin.

**Figure 2:**
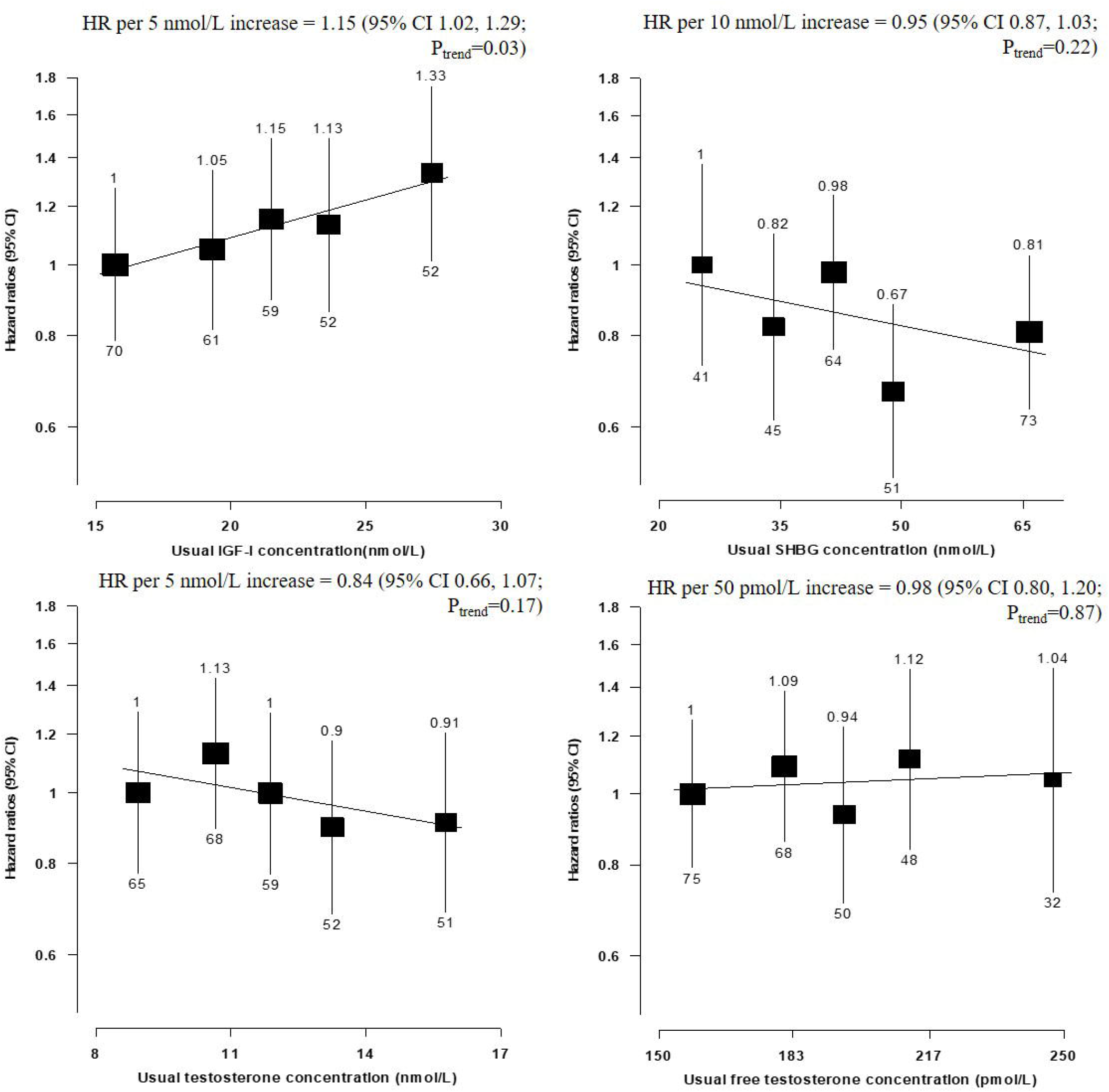
Hazard ratios of prostate cancer mortality by fifths of usual serum hormone concentrations in the UK Biobank. HRs are stratified by region (10 UK cancer registry regions) and age at recruitment (<45, 45–49, 50–54, 55–59, 60–64, and ≥65 years) and adjusted for age (underlying time variable), Townsend deprivation score (fifths, unknown), racial/ethnic group (white, mixed background, Asian, black, other, unknown), height (<170, ≥170– <175, ≥175–<180, ≥180 cm, unknown), lives with a wife or partner (no, yes), BMI (<25, ≥25–<30, ≥30–<35, ≥35 kg/m^2^), cigarette smoking (never, former, light smoker, heavy smoker, current unknown, and smoking status unknown), alcohol consumption (non-drinkers, <1-<10, ≥10-<20, ≥20 g ethanol/day, unknown), and diabetes (no, yes, and unknown). HRs for trend are adjusted for regression dilution bias. The boxes represent the HRs; the vertical lines represent the 95% CIs, with the size inversely proportional to the variance of the logarithm of the HR. The numbers above the vertical lines are point estimates for HRs, and the numbers below are the number of prostate cancer deaths. Abbreviations: BMI= body mass index; CI= confidence intervals; HR= hazard ratio; IGF-I=insulin-like growth factor-I; SHBG=sex hormone binding globulin.

Serum SHBG concentration was inversely associated with prostate cancer incidence (HR per 10 nmol/L increment=0.95, 95% CI 0.94-0.97; *P*_*trend*_<0.0001, Figure 1), but was not associated with prostate cancer mortality (Figure 2). Free testosterone was positively associated with prostate cancer incidence (HR per 50 pmol/L increment=1.10, 95% CI 1.05-1.15; *P*_*trend*_<0.0001, Figure 1). Total testosterone was not associated with prostate cancer incidence or mortality.

Risk estimates with and without adjustment for regression dilution bias are shown in Supplementary Table S2.

#### Subgroup analyses

There was no evidence of heterogeneity in the associations of IGF-I or SHBG with incident prostate cancer by any of the selected characteristics (Figures 3 and 4). There was some evidence that the inverse association between SHBG and prostate cancer incidence varied by IGF-I concentration (*P*_*he*t_=0.03); only men with lower concentrations of IGF-I (< the study median) had a reduced risk of prostate cancer (HR per 10 nmol/L increment in SHBG=0.94, 95% CI 0.92-0.97, Figure 4).

**Figure 3:**
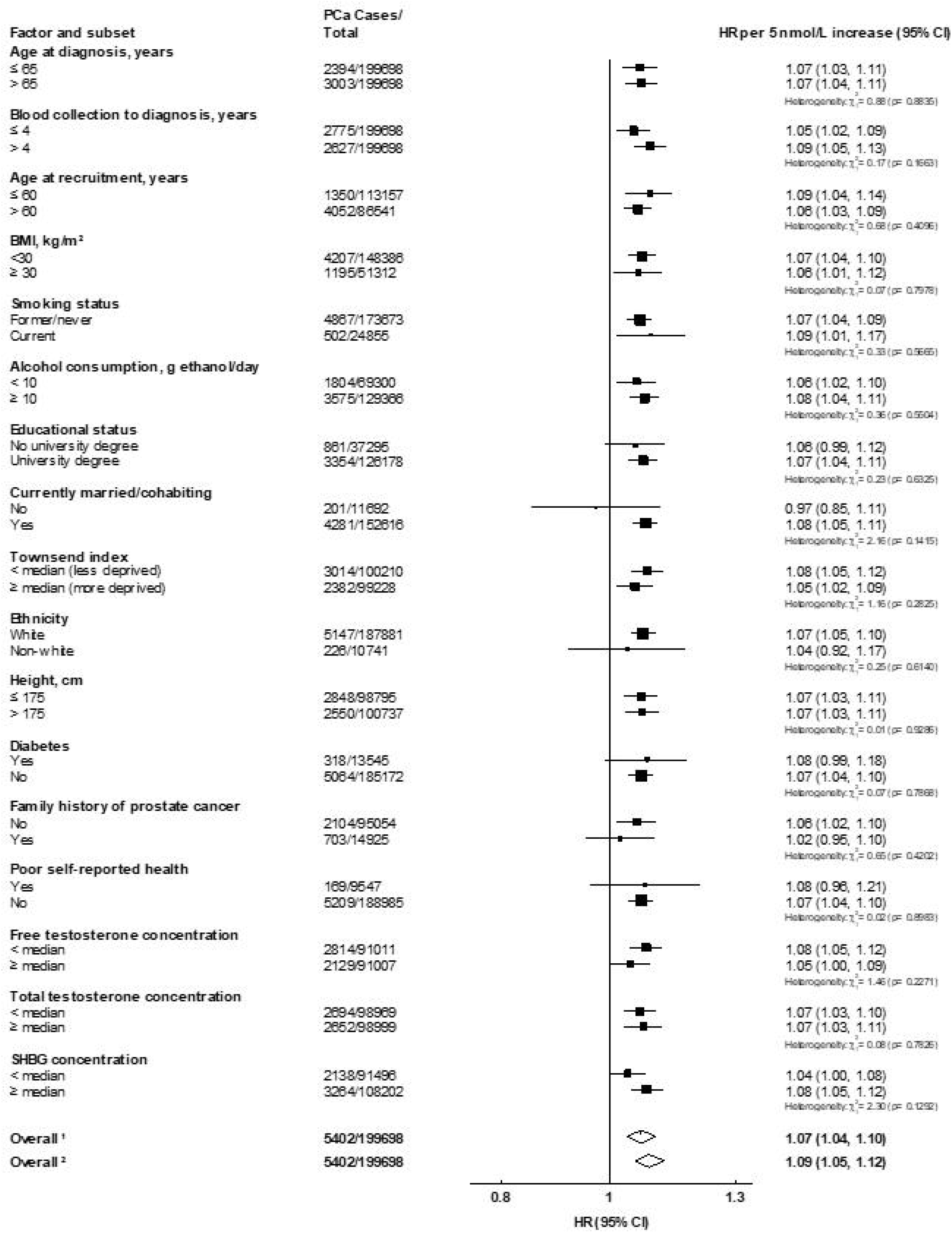
Hazard ratios of incident prostate cancer per 5 nmol/L increase in serum IGF-I concentration by subgroup in the UK Biobank. Cox models based on competing risks and compared the risk coefficients and standard errors in the two subgroups and tested using a χ^2^ test of heterogeneity. For non-case specific factors, heterogeneity was assessed using a χ^2^ interaction term. HRs are stratified by region (10 UK cancer registry regions) and age at recruitment (<45, 45–49, 50–54, 55–59, 60–64, and ≥65 years) and adjusted for age (underlying time variable), Townsend deprivation score (fifths, unknown), racial/ethnic group (white, mixed background, Asian, black, other, unknown), height (<170, ≥170– <175, ≥175–<180, ≥180 cm, unknown), lives with a wife or partner (no, yes), BMI (<25, ≥25–<30, ≥30–<35, ≥35 kg/m^2^), cigarette smoking (never, former, light smoker, heavy smoker, current unknown, and smoking status unknown), alcohol consumption (non-drinkers, <1-<10, ≥10-<20, ≥20 g ethanol/day, unknown), and diabetes (no, yes, and unknown). The boxes represent the HRs; the horizontal lines represent the 95% CIs, with the size inversely proportional to the variance of the logarithm of the HR. ^1^ Not adjusted for regression dilution bias. ^2^ Adjusted for regression dilution bias. Abbreviations: BMI= body mass index; CI= confidence intervals; HR= hazard ratio; IGF-I=insulin-like growth factor-I; PCa= prostate cancer; SD=standard deviation.

**Figure 4:**
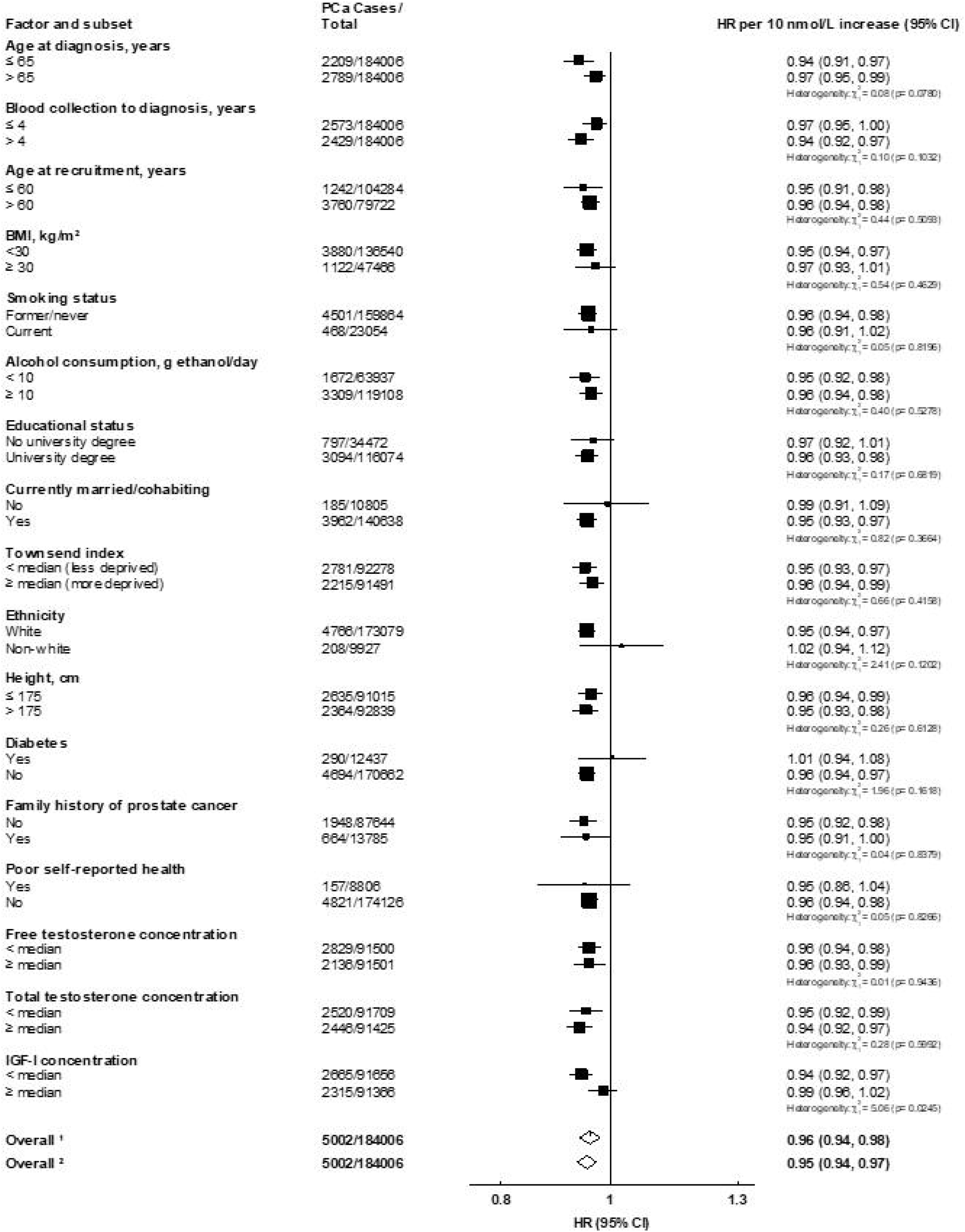
Hazard ratio of incident prostate cancer per 10 nmol/L increase in serum SHBG concentration by subgroup in the UK Biobank. Cox models based on competing risks and compared the risk coefficients and standard errors in the two subgroups and tested using a χ^2^ test of heterogeneity. For non-case specific factors, heterogeneity was assessed using a χ^2^ interaction term. HRs are stratified by region (10 UK cancer registry regions) and age at recruitment (<45, 45–49, 50–54, 55–59, 60–64, and ≥65 years) and adjusted for age (underlying time variable), and adjusted for Townsend deprivation score (fifths, unknown), racial/ethnic group (white, mixed background, Asian, black, other, unknown), height (<170, ≥170–<175, ≥175–<180, ≥180 cm, unknown), lives with a wife or partner (no, yes), BMI (<25, ≥25– <30, ≥30–<35, ≥35 kg/m^2^), cigarette smoking (never, former, light smoker, heavy smoker, current unknown, and smoking status unknown), alcohol consumption (non-drinkers, <1-<10, ≥10-<20, ≥20 g ethanol/day, unknown), and diabetes (no, yes, and unknown). The boxes represent the HRs; the horizontal lines represent the 95% CIs, with the size inversely proportional to the variance of the logarithm of the HR. ^1^ Not adjusted for regression dilution bias. ^2^ Adjusted for regression dilution bias. Abbreviations: BMI= body mass index; CI= confidence intervals; HR= hazard ratio; PCa= prostate cancer; SD=standard deviation; SHBG=sex hormone binding globulin.

There was no evidence of heterogeneity in the associations of total or free testosterone with prostate cancer incidence by any characteristics except for diabetes status. For free testosterone, there was evidence that the magnitude of the association with incident prostate cancer was greater in men who were diabetic at baseline (HR per 50 pmol/L increment=1.19, 95% CI 1.10-1.29) than in those who were not (HR=1.05, 95% CI 1.02-1.07; *P*_*het*_=0.004, Figure 5). Total testosterone was also associated with prostate cancer in men with type II diabetes, but not in men without diabetes (Supplementary Figure S2).

**Figure 5:**
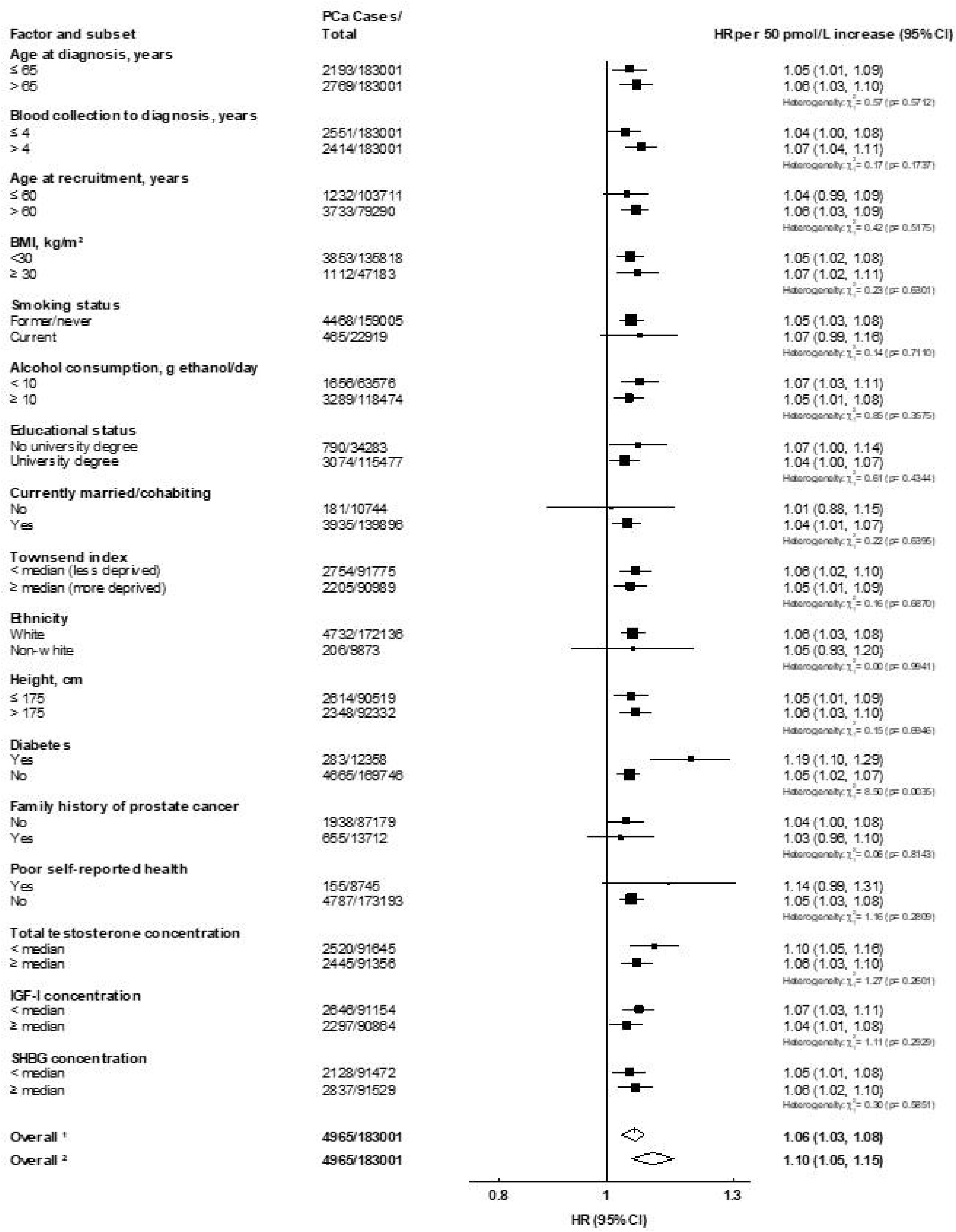
Hazard ratio of incident prostate cancer per 50 pmol/L increase in serum free testosterone concentration by subgroup in the UK Biobank. Cox models based on competing risks and compared the risk coefficients and standard errors in the two subgroups and tested using a χ^2^ test of heterogeneity. For non-case specific factors, heterogeneity was assessed using a χ^2^ interaction term. HRs are stratified by region (10 UK cancer registry regions) and age at recruitment (<45, 45–49, 50–54, 55–59, 60–64, and ≥65 years) and adjusted for age (underlying time variable), and adjusted for Townsend deprivation score (fifths, unknown), racial/ethnic group (white, mixed background, Asian, black, other, unknown), height (<170, ≥170–<175, ≥175–<180, ≥180 cm, unknown), lives with a wife or partner (no, yes), BMI (<25, ≥25– <30, ≥30–<35, ≥35 kg/m^2^), cigarette smoking (never, former, light smoker, heavy smoker, current unknown, and smoking status unknown), alcohol consumption (non-drinkers, <1-<10, ≥10-<20, ≥20 g ethanol/day, unknown), and diabetes (no, yes, and unknown). The boxes represent the HRs; the horizontal lines represent the 95% CIs, with the size inversely proportional to the variance of the logarithm of the HR. ^1^ Not adjusted for regression dilution bias. ^2^ Adjusted for regression dilution bias. Abbreviations: BMI= body mass index; CI= confidence intervals; HR= hazard ratio; PCa= prostate cancer; SD=standard deviation.

#### Further analyses

Associations with incident prostate cancer remained broadly similar when the associations were examined across tenths of the distributions (Supplementary Figure S1), and per 80 percentile increase (Supplementary Table S3). Minimally adjusted results are displayed in Supplementary Table S4. Mutual adjustment for hormones did not materially affect the risk estimates (Supplementary Table S4).

### Mendelian randomization

MR analysis using the *cis*-SNP found that IGF-I was significantly associated with a 34% increased prostate cancer risk per 5 nmol/L increment (95% CI 1.07-1.68; *P*=0.01) (Table 2).

**Table 2:**
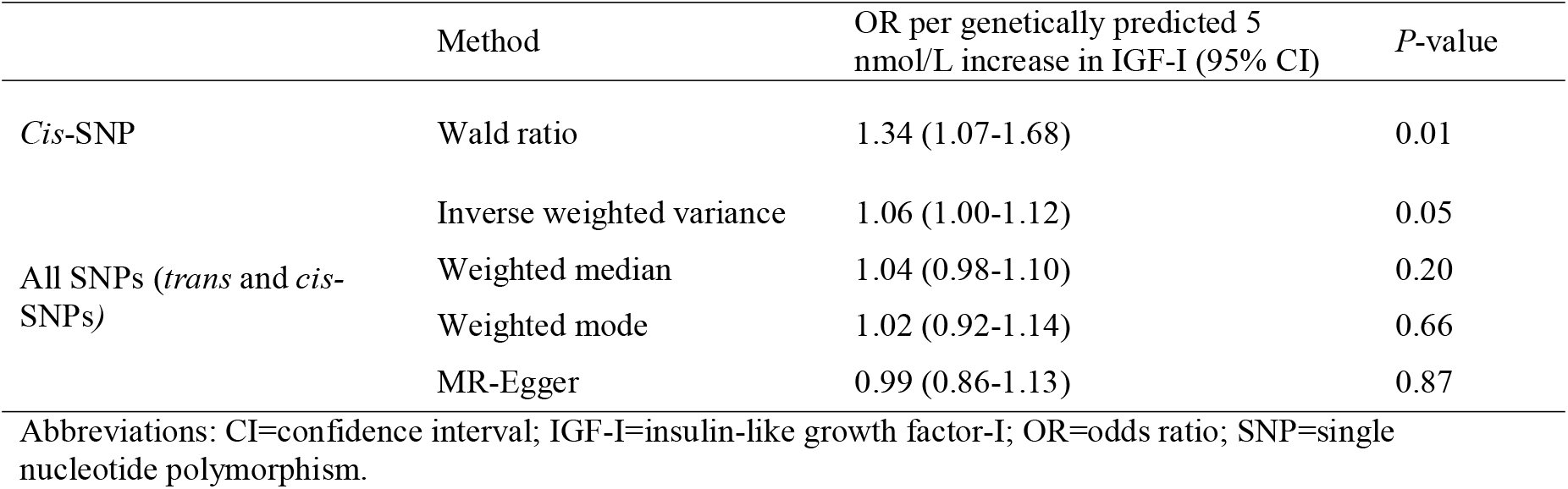
**Mendelian randomization estimates between genetically predicted circulating IGF-I concentrations and prostate cancer risk**

MR analysis including all the SNPs (both the *cis* and *trans*-SNPs) found a borderline significant association in the same direction as the *cis*-SNP results (inverse-variance weighted OR for a genetically predicted 5 nmol/L increment in IGF-I =1.06, 95% CI 1.00-1.12; *P*=0.05, Table 2). However, while the Egger intercept did not indicate the presence of directional pleiotropy, significant heterogeneity (Cochran’s Q *P*<0.0001) may have influenced standard error estimation for the inverse-variance weighted estimate (Table 2). Results were broadly consistent in leave-one-out analyses (data not shown).

PheWAS using published data showed the c*is*-SNP, rs5742653, was associated with measures of lung function and adiposity (Supplementary Table S5). There was a large amount of pleiotropy in the *trans* genetic instruments, for example, for the top 100 *trans*-SNPs most strongly associated with IGF-I there were >2,500 published genome-wide significant associations (using the PhenoScanner resource).

## Discussion

Our observational and MR analyses provide strong evidence that men with higher circulating IGF-I have an elevated risk of prostate cancer; further, our observational analyses suggest a higher risk of prostate cancer mortality in these men, suggesting that IGF-I is associated with risk for more severe forms of prostate cancer and/or may increase the risk of prostate cancer progression. Higher serum free testosterone was associated with a higher risk of prostate cancer diagnosis, which is supported by a recent MR analysis^9^. We also found that men with higher SHBG had a lower risk. Total testosterone concentration was not associated with prostate cancer incidence or mortality.

The findings of a likely causal effect of IGF-I in prostate cancer development may be due to its role in activating signalling pathways which regulate cell proliferation and apoptosis^2^. The positive relationship between IGF-I and incident prostate cancer observed is consistent with previous epidemiological evidence^4^, as well as associations observed with other cancers including breast and colorectal^35-37^. Further genetic epidemiology including fine mapping may help elucidate exactly by which mechanism variation at the *IGF1* locus associates with risk prostate and some other cancers.

Our finding of a positive association between calculated free testosterone concentration and prostate cancer diagnosis is consistent with the hypothesised importance of androgens for prostate cancer development^3^, including previous evidence from pooled nested case-control studies^8^, randomised controlled trials which aim to reduce intra-prostatic androgen signalling^38 39^ and MR analyses^9^. However, the shape of the association between circulating free testosterone and prostate cancer risk is inconsistent; here we see an approximately linear association, whereas our previous pooled analysis suggested that very low free testosterone concentrations was associated with a lower prostate cancer risk, but that risk did not change with further increments in free testosterone concentration^8^.

Previous studies have suggested that men with low free testosterone may have an increased risk of high-grade prostate tumours^8 38 39^. In this analysis prostate cancer mortality was used as a proxy of tumour aggressiveness; we did not observe an association between circulating total or free testosterone concentrations and prostate cancer mortality, although statistical power to examine this association was limited (<300 prostate cancer deaths).

Men with higher SHBG concentrations had a lower risk of prostate cancer, which is consistent with previous prospective studies^8^. MR analyses are also suggestive of an inverse relationship, but results were likely underpowered^9^. In the current study we cannot determine whether the mechanism underlying this association relates to SHBG itself, or to the action of SHBG as a carrier protein which modulates androgen access to tissues.

This analysis has several strengths. It is the largest prospective full-cohort analysis to examine hormones in relation to prostate cancer incidence and mortality. UK Biobank is a well-characterised study population and therefore we were able to adjust our risk results for a wide range of possible confounders and to investigate associations in a number of subgroups. Our results were consistent across these subgroups; in only two subgroup analyses was there weak evidence of heterogeneity in the associations with prostate cancer risk; these findings warrant further investigation, but we cannot exclude the role of chance due to multiple tests. Hormones were measured using a standardised method, therefore we were able to estimate risk associations on the absolute scale and use repeat measurements to improve the precision of risk estimates^21^. Further, by incorporating both observational and MR methods we were able to use several lines of evidence with orthogonal biases to investigate the potential causality of the associations of IGF-I with prostate cancer risk^40^. Our MR analysis of IGF-I using a *cis*-SNP is an example of the strongest case for an MR analysis, due to the strong plausibility of a biological link and a reduced likelihood of horizontal pleiotropy^29 30^, therefore the association of this *cis-*SNP indicates that IGF-I may be driving the reported associations with prostate cancer risk.

A limitation of the analysis is that prostate tumour stage and grade information are not currently available in the UK Biobank, and only incident prostate cancer data are available in the MR analyses. The UK Biobank participants are predominantly white and healthier than the sampling population, therefore selection bias may influence the results^41^ and risk estimates may not be generalizable^13^, although this is unlikely to affect the direction of the associations^42^. Relatively weak evidence from MR analyses incorporating all GWAS significant SNPs for IGF-I may reflect widespread pleiotropy for *trans*-SNPs and/or uncontrolled confounding due to ancestry^43^, which served as primary motivation to emphasise the association of the *cis* variant. Testosterone is related to other sex hormones, which have not been measured in UK Biobank, therefore associations may be at least partially be explained by other androgens, although there is little observational evidence to support this^44^. Furthermore, the predictive value of calculated free testosterone as an indicator of androgen exposure within the prostate remains under debate^45^.

PSA concentrations are partly regulated by the androgen receptor^46^; lower free testosterone concentrations may therefore reduce circulating PSA concentrations, reducing the likelihood of prostate cancer detection rather than development. Co-morbidities, socioeconomic status and poor health may affect PSA test attendance, but PSA testing attendance after baseline was not known.

In conclusion, our results implicate IGF-I and free testosterone in prostate cancer development and/or progression. This analysis of 200,000 men enabled us to quantify the associations of circulating hormones with prostate cancer risk on the absolute scale. The complementary MR for IGF-I supports a causal association. Future research will examine hormone associations by tumour stage and grade.

## Data Availability

UK Biobank data are available through a procedure described at http://www.ukbiobank.ac.uk/using-the-resource/. IGF1 GWAS data are publicly available from: http://www.nealelab.is/uk-biobank. PRACTICAL data may be available on application: http://practical.icr.ac.uk/blog/

http://www.nealelab.is/uk-biobank

http://practical.icr.ac.uk/blog/

## Abbreviations

CI: confidence interval
GWAS: genome-wide association study
HR: hazard ratio
ICD-10: International Classification of Diseases Tenth revision code 10
IGF-I: insulin like growth factor-I
LD: linkage disequilibrium
MR: Mendelian randomization
OR: odds ratio
PSA: prostate-specific antigen
SHBG: sex hormone-binding globulin
SNP: Single nucleotide polymorphism.

## Acknowledgements

This research has been conducted using the UK Biobank Resource under application number 3282 and data from the PRACTICAL consortium, CRUK, BPC3, CAPS, PEGASUS.

## Funding

This work was supported by Cancer Research UK (grant numbers C8221/A19170, C8221/A20986 and C8221/A29017). ELW was supported by the Nuffield Department of Population Health Early Career Research Fellowship. AK is supported by the Wellcome Trust (LEAP 205212/Z/16/Z). MVH works in a unit that receives funding from the UK Medical Research Council and is supported by a British Heart Foundation Intermediate Clinical Research Fellowship (FS/18/23/33512) and the National Institute for Health Research Oxford Biomedical Research Centre. RMM was supported by a Cancer Research UK (C18281/A19169) programme grant (the Integrative Cancer Epidemiology Programme) and is part of the Medical Research Council Integrative Epidemiology Unit at the University of Bristol supported by the Medical Research Council (MC_UU_12013/1, MC_UU_12013/2, and MC_UU_12013/3) and the University of Bristol. RMM is also supported by the National Institute for Health Research (NIHR) Bristol Biomedical Research Centre which is funded by the National Institute for Health Research (NIHR) and is a partnership between University Hospitals Bristol NHS Foundation Trust and the University of Bristol. KKT was funded by the Small Research Teams funding programme from the Hellenic Republic, Ministry of Education.

The Prostate cancer genome-wide association analyses are supported by the Canadian Institutes of Health Research, European Commission’s Seventh Framework Programme grant agreement n° 223175 (HEALTH-F2-2009-223175), Cancer Research UK Grants C5047/A7357, C1287/A10118, C1287/A16563, C5047/A3354, C5047/A10692, C16913/A6135, and The National Institute of Health (NIH) Cancer Post-Cancer GWAS initiative grant: No. 1 U19 CA 148537-01 (the GAME-ON initiative).

We would also like to thank the following for funding support: The Institute of Cancer Research and The Everyman Campaign, The Prostate Cancer Research Foundation, Prostate Research Campaign UK (now PCUK), The Orchid Cancer Appeal, Rosetrees Trust, The National Cancer Research Network UK, The National Cancer Research Institute (NCRI) UK. We are grateful for support of NIHR funding to the NIHR Biomedical Research Centre at The Institute of Cancer Research and The Royal Marsden NHS Foundation Trust.

The Prostate Cancer Program of Cancer Council Victoria also acknowledge grant support from The National Health and Medical Research Council, Australia (126402, 209057, 251533,, 396414, 450104, 504700, 504702, 504715, 623204, 940394, 614296,), VicHealth, Cancer Council Victoria, The Prostate Cancer Foundation of Australia, The Whitten Foundation, PricewaterhouseCoopers, and Tattersall’s. EAO, DMK, and EMK acknowledge the Intramural Program of the National Human Genome Research Institute for their support.

Genotyping of the OncoArray was funded by the US National Institutes of Health (NIH) [U19 CA 148537 for ELucidating Loci Involved in Prostate cancer SuscEptibility (ELLIPSE) project and X01HG007492 to the Center for Inherited Disease Research (CIDR) under contract number HHSN268201200008I] and by Cancer Research UK grant A8197/A16565. Additional analytic support was provided by NIH NCI U01 CA188392 (PI: Schumacher).

Funding for the iCOGS infrastructure came from: the European Community’s Seventh Framework Programme under grant agreement n° 223175 (HEALTH-F2-2009-223175) (COGS), Cancer Research UK (C1287/A10118, C1287/A 10710, C12292/A11174, C1281/A12014, C5047/A8384, C5047/A15007, C5047/A10692, C8197/A16565), the National Institutes of Health (CA128978) and Post-Cancer GWAS initiative (1U19 CA148537, 1U19 CA148065 and 1U19 CA148112 – the GAME-ON initiative), the Department of Defence (W81XWH-10-1-0341), the Canadian Institutes of Health Research (CIHR) for the CIHR Team in Familial Risks of Breast Cancer, Komen Foundation for the Cure, the Breast Cancer Research Foundation, and the Ovarian Cancer Research Fund.

The BPC3 was supported by the U.S. National Institutes of Health, National Cancer Institute (cooperative agreements U01-CA98233 to D.J.H., U01-CA98710 to S.M.G., U01-CA98216 to E.R., and U01-CA98758 to B.E.H., and Intramural Research Program of NIH/National Cancer Institute, Division of Cancer Epidemiology and Genetics).

CAPS GWAS study was supported by the Swedish Cancer Foundation (grant no 09-0677, 11-484, 12-823), the Cancer Risk Prediction Center (CRisP; www.crispcenter.org), a Linneus Centre (Contract ID 70867902) financed by the Swedish Research Council, Swedish Research Council (grant no K2010-70X-20430-04-3, 2014-2269)

PEGASUS was supported by the Intramural Research Program, Division of Cancer Epidemiology and Genetics, National Cancer Institute, National Institutes of Health.

## Disclosure

The authors have no conflicts of interest to disclose.

## Disclaimers

Department of Health and Social Care disclaimer: The views expressed are those of the author(s) and not necessarily those of the NHS, the NIHR or the Department of Health and Social Care.

Where authors are identified as personnel of the International Agency for Research on Cancer/ World Health Organization, the authors alone are responsible for the views expressed in this article and they do not necessarily represent the decisions, policy or views of the International Agency for Research on Cancer / World Health Organization.

## Notes

### Competing Interest Statement

The authors have declared no competing interest.

